# First experiences of whole-head OP-MEG recordings from a patient with Epilepsy

**DOI:** 10.1101/2021.09.28.21264047

**Authors:** Stephanie Mellor, Umesh Vivekananda, George C. O’Neill, Tim M. Tierney, David Doig, Robert A. Seymour, Nicholas Alexander, Matthew C. Walker, Gareth R. Barnes

## Abstract

Optically Pumped Magnetometer based Magnetoencephalography (OP-MEG) has significant potential for clinical use in pre-surgical planning in epilepsy. Unlike current clinical MEG, the sensors do not require cryogenic cooling and so can be placed directly on the patient’s head. This allows the patient to move during the recording and means that the sensor positions can be chosen to suit the patient’s head shape and suspected epileptogenic focus. However, OP-MEG is a new technology, and more work is needed to demonstrate this potential. We present OP-MEG recordings from a patient (male in their 30s) with a radiologically identifiable focal cortical dysplasia (FCD) in the right superior frontal sulcus approximately 1.9 cm^3^ in volume. Previous scalp EEG studies and prolonged video-EEG telemetry did not identify any interictal epileptiform abnormalities. We recorded 30 minutes of OP-MEG with 62-channel, whole-head sensor coverage. During the experiment, the patient’s head was unconstrained. We localised interictal epileptiform discharges (IEDs) from this recording with a beamformer and by fitting a dipole to the averaged IED data. Both the beamformer peak and dipole fit locations were within 2.3 cm of the MRI lesion boundary. This single subject, proof-of-concept recording provides further evidence that OP-MEG can be a useful and minimally invasive tool in the clinical evaluation of epilepsy.

## 1 Introduction

Magnetoencephalography (MEG) can be a useful imaging modality for pre-surgical planning in epilepsy (Knowlton, 2006; Sutherling *et al*., 2008; Englot *et al*., 2015; Murakami *et al*., 2016; Ramanujam *et al*., 2017). MEG can guide the implantation of intracranial electrodes (Sutherling *et al*., 2008), map the eloquent cortex (Stufflebeam, Tanaka and Ahlfors, 2009; Collinge *et al*., 2017) and predict seizure freedom after surgery (Englot *et al*., 2015). In a recent retrospective study of 1000 patients (Rampp et al., 2019), it was shown that complete resection of MEG localisations was associated with significantly higher probability of Engel 1 outcome (free from disabling seizures) over the 10 years proceeding surgery.

However, MEG is often not clinically available. A survey of epilepsy surgery centres found only 1/3 of the centres had access to MEG (Mouthaan *et al*., 2016). Optically Pumped magnetometer-based MEG (OP-MEG) has many advantages which could make it more accessible in a clinical setting than the currently more standard superconducting quantum interference device (SQUID) based MEG. The sensors do not need to be cryogenically cooled and can be worn directly on the head, meaning that there is scope for the patient to move. This is particularly important for less compliant patient groups such as children (Wehner *et al*., 2008; Larson and Taulu, 2017) or patients requiring long-term monitoring for the purpose of capturing ictal events. Additionally, the sensors can be placed directly on the scalp, rather than the one-size-fits-all array which is typical for SQUID-MEG. This means that it is possible to capture maximal signal for any head-size. This can increase the SNR of observed interictal epileptiform discharges (Feys *et al*., 2021) and it has been shown with high-T_C_ on-scalp SQUID-MEG, that higher proximity of the sensors to the scalp can increase the number of interictal discharges that are observed (Westin *et al*., 2020). This improvement would be particularly important for children, due to their smaller head size (Boto *et al*., 2016; Hill *et al*., 2019) and is pertinent for epilepsy since there is an incidence peak in childhood (Kotsopoulos *et al*., 2002; Fiest *et al*., 2017).

Previous OP-MEG studies have demonstrated epileptogenic discharges in mice (Alem *et al*., 2014), in a single subject (Vivekananda *et al*., 2020) and in 5 pediatric patients (Feys *et al*., 2021). One of the main applications for OP-MEG in epilepsy is identification of the epileptogenic focus for pre-surgical planning. In this study, we report the results of whole-head OP-MEG from a single patient with focal cortical dysplasia (FCD) in the right superior frontal sulcus. As the patient has a radiologically identifiable lesion, we have a prior expectation of the epileptogenic focus. This proof-of-concept experiment shows that further studies with more patients would be worthwhile and could provide a viable pathway to the clinical translation of OP-MEG.

## 2 Methods

A male in their 30s with FCD in the right superior frontal sulcus participated in this study. He is currently on four antiepileptic agents and experiences up to six seizures a night. Previous interictal scalp EEG and video-EEG telemetry had not recorded any interictal epileptiform activity. Seizures had been recorded and semiology was consistent with seizures originating from the area of dysplasia, but the ictal EEG was obscured by artefact. Ethical approval for the study was granted by the Medicines and Healthcare products Regulatory Agency, and informed consent was obtained from the patient prior to participation. Throughout the experiment, the participant was monitored by a clinician from outside of the recording room via video and audio.

### 2.1 Recordings

30 minutes of MEG data was recorded from 37 QuSpin Gen-2 QZFM OPMs. The sensors were placed into a scanner-cast bespoke to the participant (Boto *et al*., 2017). This scanner-cast was 3D-printed to match their scalp surface, as measured from a previous structural 3T MRI (Meyer *et al*., 2017). The sensors were positioned approximately evenly around the head, with greater coverage over the right, frontal cortex, as shown in Figure 1. The magnetic field perturbations along two axes of the OPMs – one radial to the scalp and one tangential – were recorded, meaning that there were 74 recording channels in total. 4 OPMs were later removed from the analysis due to abnormally high noise floors and 2 were removed due to faults in the OPM heating hardware creating erroneous signals. This left 62 recording channels.

**Figure 1.**
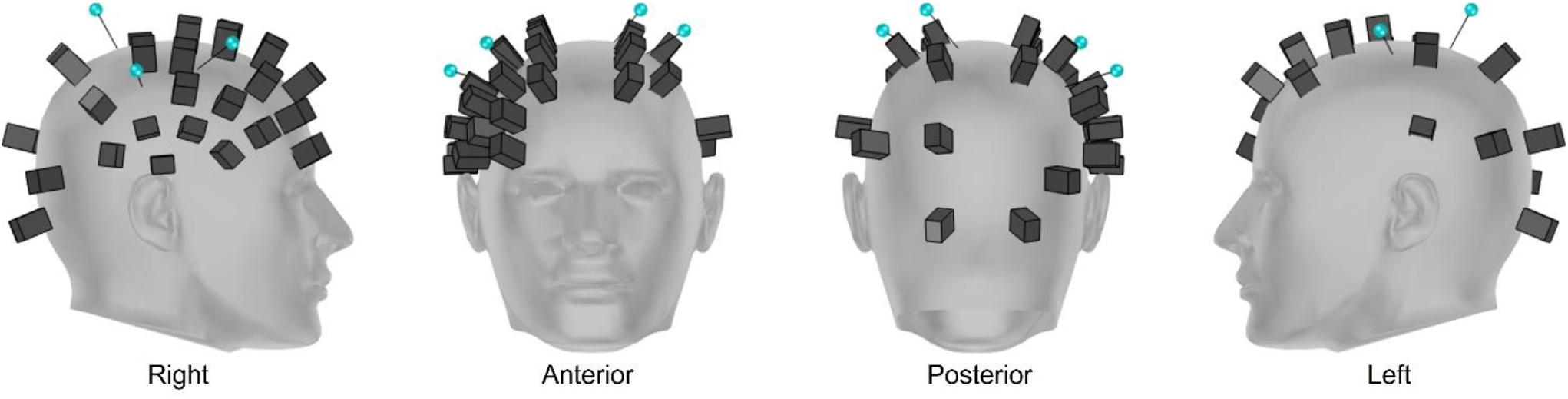
Sensor and marker positions for the OP-MEG recordings. The black cubes represent the OPMs. The blue dots are the retro-reflective markers for the motion tracking. Although the scanner-cast was subject specific, the OPM positions are shown in MNI coordinates and the head mesh displayed is a generic head object file (https://www.turbosquid.com/3d-models/male-head-obj/346686). Only the 31 OPMs used for analysis are shown.

The recording was performed in a 4-layer Magnetically Shielded Room (MSR) (Magnetic Shields, Ltd.; internal dimensions 3 m × 4 m × 2.2 m). The participant was seated on a beanbag for the experiment and, consequently, remained within the central cubic meter of the room. The static field at the centre of the UCL room is approximately 2 nT and the field gradient is approximately 1 nT per metre (Mellor *et al*., 2021). No additional active shielding was used. The recording was split into three runs of 10 minutes each. In the first and second runs, the patient was reading a book. In the second run, for the first and last minute of the 10-minute recording, we asked the participant to move their head. In the third and final run, the participant sat still and at rest with their eyes open. No overt task was performed during the recording.

The OPM data was recorded using a custom acquisition software, built in Labview. Each OPM channel was measured as a voltage (±5 V, 500 Hz antialiasing hardware filter) and sampled at 6 kHz by a National Instruments (NI) analogue-to-digital converter (NI-9205, 16-bit, ± 10 V input range) using QuSpin’s adapter (https://quspin.com/products-qzfm/ni-9205-data-acquisition-unit/). This voltage was then multiplied by a calibration factor to calculate the recorded magnetic field.

In addition to the OPM data, the patient’s movements were recorded with an array of 6 OptiTrack Flex 13 cameras. The cameras were positioned in three corners of the room, with one low and one close to the ceiling in each corner. All cameras were positioned so that the participant’s head was within their field of view. The cameras were calibrated according to the manufacturer’s instructions, using the OptiTrack CW-500 calibration wand. To track the participant’s head, four retroreflective markers were placed onto the OPM scanner-cast. These are shown in blue in Figure 1. A rigid body was created from the four markers in Motive, the acquisition programme associated with the OptiTrack cameras. The rigid body position and orientation was recorded by the Flex 13 motion tracking cameras and then the corresponding positions and orientations of the OPM channels were calculated at each time point.

The marker positions were recorded at 120 Hz. A 5 V pulse was used to synchronize the motion tracking with the MEG recordings. Where markers were occluded, usually due to the wires from the OPMs, the missing data was filled in using Motive. Firstly, for any gap where only one marker was missing, a pattern-based method was used to fill in the missing marker data. In this case, the other three markers are used to estimate the position of the missing one. Then for the remaining gaps where multiple markers were occluded, cubic spline interpolation was used to fill in the missing data.

### 2.2 Analysis

Before IED detection, we performed the following pre-processing steps. Firstly, the OPM data was downsampled to 240 Hz using SPM12. The start and end were then trimmed to match the motion tracking recording and the motion data was upsampled to 240 Hz using linear interpolation in order to match the OPM data. A 10 Hz, 6th order Butterworth low-pass filter was applied to the rigid body position and orientation using FieldTrip to remove high frequency noise caused by marker vibrations.

Three 5th order Butterworth notch filters were applied to the MEG data using FieldTrip. One was at 50 Hz to remove the mains noise; the other two were at 37 Hz and 83 Hz to remove noise caused by the motion tracking cameras. Environmental noise was then reduced by modelling the background magnetic field as a homogeneous field at each timepoint (Tierney *et al*., 2021). A 5th order Butterworth high-pass filter was then applied to the MEG data at 1 Hz.

IEDs were identified by visual inspection of the pre-processed data. IEDs were identified and manually marked on the OP-MEG traces for each session by consensus between a researcher (SM) and an experienced clinician (UV). The reviewers were blinded to the magnetometer positions. These selections were then used to create 2 s trials. The centre of the epileptiform trials was determined by maximising the cross-correlation with the fourth identified IED trial. For each IED trial, the central timepoint was shifted by ±0.5 s in steps of 1/240 s and the mean (across channels) correlation between the shifted trial and fourth IED trial was calculated. The shift which corresponded to the maximum correlation was chosen to be the centre of the IED trial. The remaining data was split into 2 s baseline trials. To ensure that the identified epileptiform activity was not caused by movement artefact, we set a threshold for the amount of variance in the OPM data in the trial which could be explained by the patient’s position and orientation, above which a trial was rejected. We set this threshold based on runs 1 and 3, since there was more movement in run 2 which made it unrepresentative of the other two recordings. The threshold was set so that 20% of trials in runs 1 and 3 were rejected. This equated to 13.8% of the variance in the trial and led to rejection of 43% of the data, leaving 11 good epileptiform trials and 500 baseline trials. The implications of this conservative rejection criteria are considered further in the discussion.

Source localisation was performed using an F contrast on the source power output of an LCMV beamformer from DAiSS (https://github.com/spm/DAiSS). The epileptiform activity was compared with other sections of the data where no abnormal or epileptiform activity was identified. A confidence volume was obtained by bootstrapping which trials were used in the source localisation and then looking at the global peak location for 100 different trial selections. We also performed a dipole fit of the average spike peak (between 0.9 s and 1.1 s in the averaged trial) in FieldTrip. In all cases, we used the Nolte single shell head model (Nolte, 2003).

## 3 Results

In total, the data was split into 893, 2 s trials, 382 of which were rejected due to the degree of the data which was explained by movement in the trial. The percentage of variance in the OPM data which is explained by the rotation and position of the patient’s head in each trial is shown in Figure 2A. Encouragingly, for most trials, less than 20% of the data can be explained by movement. The distance travelled by the patient in each accepted 2 s trial is shown in Figure 2B. This suggests that these trials were accepted because there is little movement in them.

**Figure 2.**
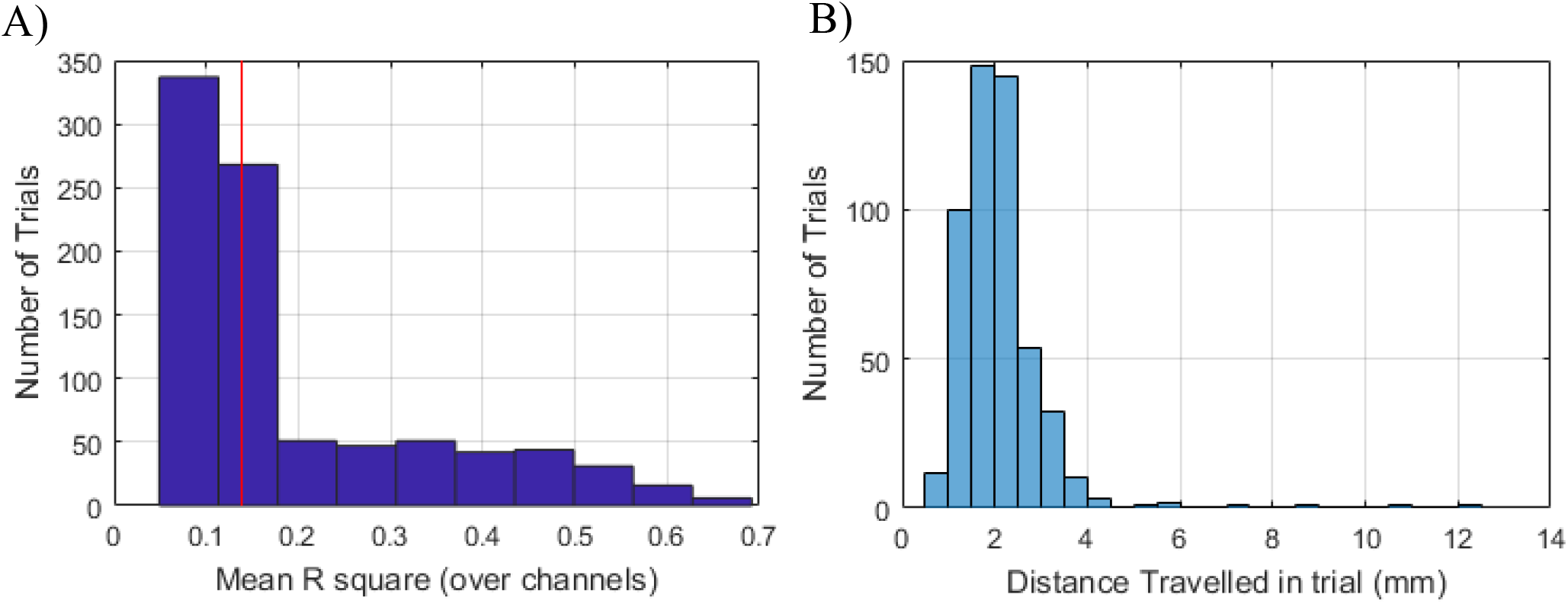
A) Histogram of the coefficient of determination (***R***^**2**^) when using the position and orientation of the head to explain the OPM data for each trial. The red line indicates the threshold above which trials were rejected. B) The degree of movement, quantified by the maximum distance travelled, in the accepted trials.

Figure 3 shows the sensor level results of the accepted spike trials. In Figure 3A, a single spike trial is shown on all channels. The selected trial is shaded in blue. Each individual spike trial is shown in Figure 3B for a single channel. The average of all 11 trials is also shown. The topography of the average is shown in Figure 3C for the radially oriented OPM channels.

**Figure 3.**
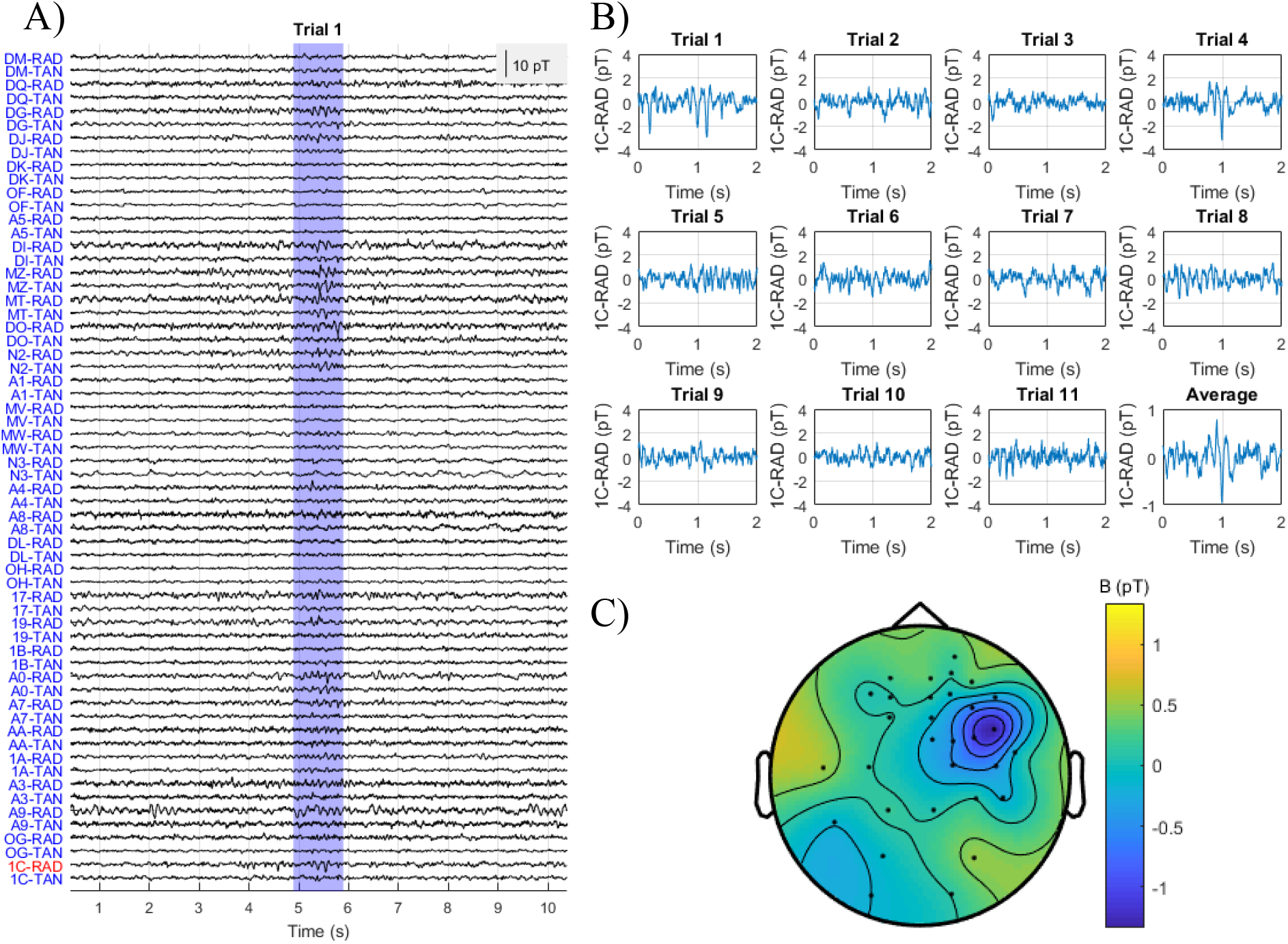
A) The sensor level data for a 10 s segment of the OPM data around trial 4. The labels along the left side are the names of the different OPM channels. The first two letters indicate the OPM, while the end of the name (RAD or TAN) indicates whether the channel is radial (RAD) or tangential (TAN) to the patient’s scalp. The selected spike trial is shaded in blue. B) Each of the individual IED trials only on channel 1C-RAD. The average is also shown. C) Topography of the average IED.

The source localisation from this epileptiform activity is shown in Figure 4. For both localisation methods, the dysplasia has been circled where visible. The results in Figure 4A are from a beamformer F contrast between epileptiform and non-epileptiform trials. A 2 mm volumetric grid was used for the possible source locations. The peak F-value is shown by the crosshairs. The F-statistics shown have been thresholded at 50% of the maximum. The localisation from OP-MEG lies on the right superior frontal gyrus, while in MRI, the lesion appears in the sulcus. The global peak of the beamformer is at (40.51, -6.53, 66.95) in MNI coordinates, which equates to 2.32 cm from the centre of the identified MRI lesion location ((20.12, -16.87, 58.94) in MNI coordinates), and 1.81 cm from the lesion boundary.

**Figure 4.**
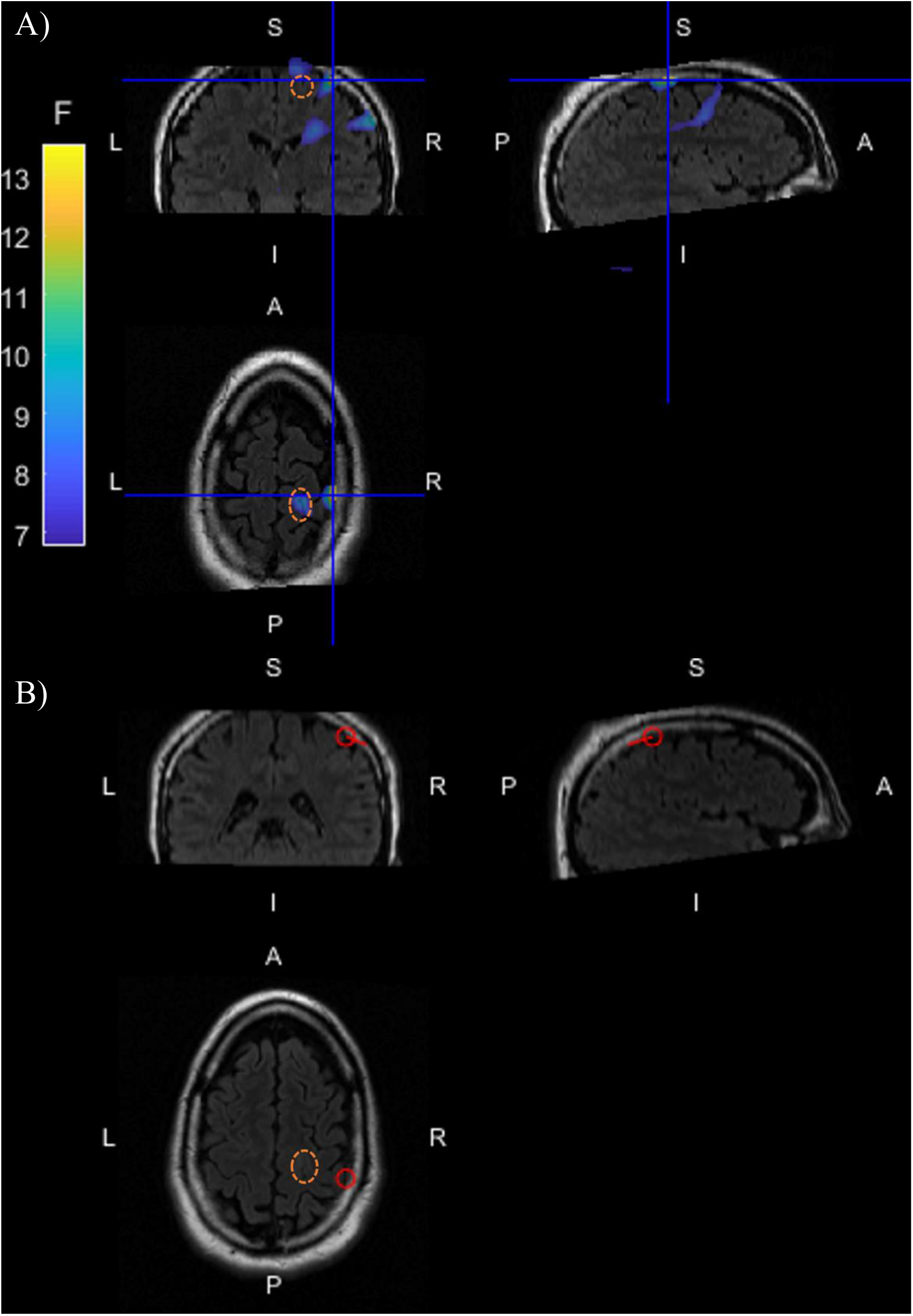
Source localisation of epileptiform activity identified in OP-MEG. The dysplasia has been circled with a dashed orange line where visible. A) Multivariate LCMV beamformer localisation of the epileptiform activity. The colour indicates the F-statistic for a comparison of epileptiform activity > baseline. The cross-hairs mark the global maximum F-value. B) Dipole fit of the average spike activity. The red ring marks the found dipole location; the line is its orientation.

Figure 4B shows the dipole fit of the average IED, shown at the scalp level in Figure 3C. Like the beamformer, the location lies on a gyrus close to the sulcus containing the dysplasia. The location ((47.46, -24.43, 60.7) in MNI coordinates) is more posterior than the beamformer global peak and is 2.74 cm from the centre of the dysplasia as identified in MRI, and 2.27 cm from the lesion boundary.

We then took the beamformer results in two directions. Firstly, we constructed virtual electrodes across a grid to look at the time series of this beamformed activity. This is shown in Figure 5. The time series in Figure 5 corresponds to the time around the first identified IED, as shown at the OPM sensor level in Figure 3A. The spike, as identified in the OP-MEG data, is clearly visible. Constructing virtual electrodes in this way, either inside the head or on the scalp, could be one way to reduce the impact of interference in OP-MEG (Seymour *et al*., 2021).

**Figure 5.**
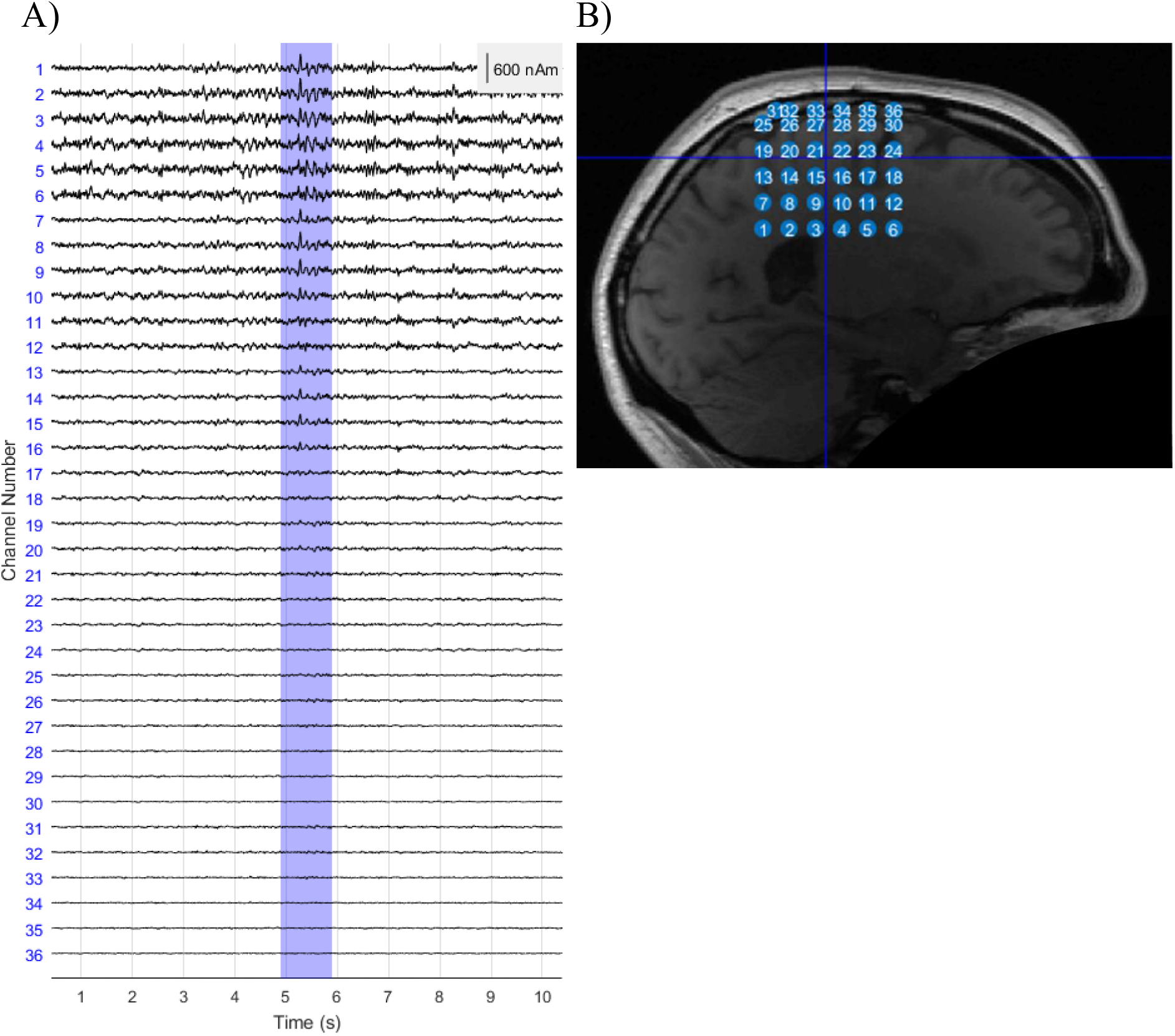
Virtual electrode time series along a 1 cm-spaced grid around the MRI FCD location. A) The time series of each virtual electrode for spike trial 1. The channel numbers correspond to the positions labelled in B).

We also bootstrapped the beamformer, in order to give a confidence volume for the localisation. We looked at 50 different bootstrap re-samplings of the OP-MEG trials and localised each one. We then looked at the global peak location for each localisation. The F values of the peaks ranged from 12.7 to 14.4 with a mean of 13.5 ± 0.1. The locations were also highly consistent across samples, with a mean in the superior frontal gyrus, 2.39±0.01 cm from the centre of the MRI FCD, 1.86±0.01 cm from the boundary. The 95% confidence volume was less than 8 mm^3^, the grid spacing of the possible source locations.

## 4 Discussion

We report whole-head OP-MEG recordings of epileptiform activity. This patient had an anatomically identifiable FCD and our source localisation is consistent with epileptiform activity arising from a region bordering the FCD. The patient was seated and head-movement was unconstrained. This is positive for future studies involving recordings over long time periods or with cohorts who are less able to remain still.

We have argued here that MRI-lesion positive is a sensible patient population to evaluate OP-MEG, since the lesion location is known. The lesion is a clear anatomical marker and it is highly likely that epileptiform activity arises from either within or around this highly epileptogenic lesion (Jackson, Kuzniecky and Berkovic, 2005). Findings from other imaging modalities have shown, however, that the seizure generating zone may extend beyond the structural lesion seen in MRI (Aubert *et al*., 2009). This provides a certain degree of confidence that our OP-MEG recordings could be useful for delineating the epileptogenic zone and supports the need for studies in larger numbers of patients with surgical outcome data. Additionally, and reassuringly, Feys *et al*., (2021) have shown a direct and compelling comparison between OPM and SQUID recordings in the estimated epileptogenic zone in 5 children with epilepsy.

The study had a number of limitations. IED detection was made more difficult by movement artefacts and other background noise from, for example, cars on the road outside of the OP-MEG shielded room. This is arguably more of a problem for OP-MEG than for SQUID-MEG since SQUID-MEG typically uses gradiometers which provide inherent noise reduction (Hämäläinen *et al*., 1993). While there are gradiometer OP-MEG designs (Nardelli *et al*., 2020; Zhang *et al*., 2020), our array was based on magnetometers. It has been shown that high SNR OP-MEG data can be achieved even when there is large participant movement (Boto *et al*., 2018; Seymour *et al*., 2021), but in these scenarios, the timing of the trials is set by an external stimulus. Here however, we required a researcher or clinician to manually select the interesting events in the sensor level data; sudden head-movements or movements of cables had a spike-like appearance which made this selection difficult. We therefore used a conservative criterion to reject epochs showing movement related activity. Consequently, whilst we did not constrain head movement in this study, we have not analyzed data during the movement itself.

There are hardware solutions to reduce these issues. In this study, the most pernicious noise was due to the OPM cables moving relative to one another. This produces artefacts which can appear as opposing deflections on nearby sensors. This can be reduced by improving the shielding on the cables and with better cable management on the scanner-cast. Additionally, while it is not ideal for comfort and suitability for certain populations, cable and movement noise can be minimized with alternative OP-MEG set-ups which keep the sensors stationary (Johnson, Schwindt and Weisend, 2013; Boto *et al*., 2017; Iivanainen *et al*., 2019; Limes *et al*., 2020; Vivekananda *et al*., 2020). Other background magnetic noise, from for example urban traffic or the underground train system, can also be reduced with additional, external, active magnetic shielding (Holmes *et al*., 2019, 2021; Iivanainen *et al*., 2019; Zhang *et al*., 2020; Rea *et al*., 2021). We were fortunate here to have an excellent MSR and so chose not to use additional external coils, to maximize the space available to the patient. Alternatively, the challenges in IED detection could motivate a sparse, simultaneous EEG recording (Boto *et al*., 2019), since it is already well studied and could provide this necessary ground truth.

There are also multiple software-based approaches to minimising OP-MEG noise. Here we have modelled the background field as a homogeneous field across the sensors and corrected for the model predictions (Tierney *et al*., 2021). Alternative or additional approaches include signal space separation (SSS) (Taulu and Kajola, 2005), synthetic gradiometry (Fife *et al*., 1999; Boto *et al*., 2016), signal-space projection (SSP) (Uusitalo and Ilmoniemi, 1997) and using the participant’s movements to model the static field within the room (Mellor *et al*., 2021). Beamformers have also been used to reduce components of MEG data which do not originate from the brain (Cheyne *et al*., 2007; Van Klink, Hillebrand and Zijlmans, 2016; Seymour *et al*., 2021). Virtual sensors constructed from the output of a beamformer, as we showed in Figure 5, could foreseeably be a sensible way to present OP-MEG data. This would reduce interference and improve the probability of detecting more subtle epileptiform activity (Van Klink, Hillebrand and Zijlmans, 2016). This is already used in epilepsy in synthetic aperture magnetometry and excess kurtosis mapping (SAM(g2)), in which a beamformer is used to create virtual electrodes in a grid across the cortex. The excess kurtosis in each virtual electrode is calculated and mapped (Robinson *et al*., 2004). For pediatric epilepsy surgery in particular, this pipeline has been shown to be predictive of seizure outcome (Gofshteyn *et al*., 2019). As such, there is still much scope for research into how best to process and present OP-MEG data for epilepsy.

## 5 Conclusion

Here we have presented whole-head OP-MEG recordings from a patient with epilepsy. The localisation of the observed interictal activity was within 2.3 cm of the boundary of the FCD lesion, as found in MRI, and appropriately located for the patient’s epilepsy for both beamformer and dipole localisations. The patient was seated and their head was unconstrained, which is encouraging for future studies with cohorts who are less able to remain still and for studies over long time periods.

## Data Availability

Anonymized OP-MEG data would be available on reasonable request from the corresponding author.

## 7 Author Contributions

SM, UV, GON, TT, RS, NA, MW and GB contributed to the conception and design of the study. SM, UV and GON recorded the data presented here. SM analysed the data. UV and DD contributed to the interpretation of the results. SM wrote the first draft of the manuscript. All authors contributed to manuscript revision, read and approved the submitted version.

## 9 Acknowledgments

The authors would like to thank Vladimir Litvak, Sven Bestmann and Eleanor Maguire for their helpful discussions and ongoing support. We would like to thank Mark Lim and Chalk Studios for designing and making the scanner-cast used in this experiment. Thanks also to Vishal Shah and QuSpin, David Woolger and Magnetic Shields Limited and our collaborators at the University of Nottingham for their continued support.

